# Breath biomarkers of insulin resistance in pre-diabetic Hispanic adolescents with obesity

**DOI:** 10.1101/2021.06.23.21259399

**Authors:** Mohammad S. Khan, Suzanna Cuda, Genesio M. Karere, Laura Cox, Andrew C. Bishop

## Abstract

**Background:** Insulin Resistance (IR) affects a quarter of the world’s adult population and is a major factor in the pathogenesis of cardio-metabolic disease. Non-invasive sampling of exhaled breath contains metabolic markers indicative of underlying systemic metabolic abnormality.

**Method:** In this pilot study, we implemented a non-invasive breathomics approach, combined with random forest machine learning, to investigate metabolic markers from pre-diabetic Hispanic adolescents with obesity as indicators of abnormal metabolic regulation.

**Findings:** Exhaled breath collection using the ReCIVA breathalyzer is feasible in an adolescent population. We have identified a signature of breath metabolites (breath-IR model) which correlates with Homeostatic Model Assessment of Insulin Resistance (HOMA-IR) (R = 0.95, *p* <0.001). A strong correlation was also observed between the breath-IR model and the blood glycemic profile (fasting insulin R=0.91, *p* <0.001 and fasting glucose R=0.80, *p* <0.001). Among tentatively identified metabolites, limonene, undecane, and 2,7-dimethyl-undecane, significantly cluster individuals based on HOMA-IR (*p* =0.003, *p* =0.002, and *p*<0.001, respectively). Our breath-IR model differentiates between adolescents with and without IR with an area under the receiver operating characteristic curve of 0.87, after cross-validation.

**Interpretation:** Identification of a breath metabolite signature indicative of IR in prediabetic Hispanic adolescents with obesity provides evidence of the utility of exhaled breath metabolomics for assessing systemic metabolic dysregulation. A simple and non-invasive breath-based test has utility as a diagnostic tool for monitoring IR progression, potentially allowing for earlier detection of IR and implementation of early interventions to prevent onset of type 2 diabetes mellitus.

**Funding:** This study was funded by The Healthy Babies Project, Texas Biomedical Research Institute, San Antonio, TX.

## 1. INTRODUCTION

An estimated 1.26 billion adults worldwide have insulin resistance (IR), a complex pathophysiological state connected to an imbalance between insulin and glucose metabolism, which is strongly associated with the development of cardio-metabolic disease^1-3^. Although aging is considered a strong predictor of IR, a meta-analysis of 18 population-based studies from 13 countries indicated that approximately 312 million children globally have IR^4^, suggesting that other factors also contribute to IR development earlier in life. One major contributing factors is obesity and in the United States between 2015 and 2016, 18.5% of youth between the ages of 2-19 were obese^5,6^. Severe obesity is also racially and ethnically disproportionate with two-to-four fold higher rates among African American or Hispanic populations compared to their Caucasian counterparts^6^, therefore more research is needed to investigate these populations to slow or prevent long term health consequences.

The exact pathophysiology of IR is unclear but involves multiple associations of cellular dysfunction. IR is inversely related to the insulin sensitivity in insulin-dependent cells such as skeletal muscle, adipocytes, and cardiomyocytes^7^. When insulin sensitivity is low, these cells fail to respond to insulin signaling transduction, which is required for glucose uptake^8^. As a result, IR is a common feature in multiple metabolic disorders including obesity, dyslipidemia, hypertension, atherosclerosis, nonalcoholic fatty liver disease (NAFLD), type 2 diabetes mellitus (T2DM), and some cases of type 1 diabetes mellitus (T1DM)^1,8,9^. One potential candidate promoting IR is obesity-induced inflammation, as many cytokines and inflammatory mediators, particularly tumor necrosis factor‐α (TNF‐α), monocyte chemotactic protein‐1 (MCP‐1), C‐reactive protein (CRP), and interleukins are upregulated in individuals with IR^10^. Another factor that plays an important role in IR is reactive oxygen species (ROS). Individuals with IR have reduced antioxidants and increased production of the ROS leading to increased systemic oxidative stress^11^. ROS induced oxidative stress metabolites such as hydrogen peroxide, protein carbonyls, protein oxidative products, carbohydrate metabolites, short chain aldehydes, ketones, and hydrocarbons have been found to strongly associated with IR^11^. Further research into these factors associated with IR development at the earlier stages or prior to the onset of clinical symptoms may uncover early biomarkers of IR. The addition of these biomarkers into the clinical setting would add to the clinical tools available for clinicians to identify at risk individuals prior to onset of disease and initiate earlier interventions.

Current clinical practice for assessing IR utilizes blood-based measures requiring fasting for 8 h and multiple visits to a medical facility for blood sampling and consultation. This is a burden for individuals who may benefit from frequently monitoring IR status, particularly young children and adolescents who rely on parents or guardians to facilitate this medical care. Individuals with IR may not have blood glucose levels that are in a range to be clinically diagnosed as T2DM but may be in a pre-diabetic stage of metabolic dysfunction. This stage is particularly important because it is a window of time when health and weight management strategies can be implemented to improve long-term cardio-metabolic health outcomes. Evidence suggests 70% of IR individuals will develop cardio-metabolic disease within 15-20 years^12,13^ without any intervention. This emphasizes the need to develop clinically relevant diagnostics tools to identify at risk individuals for intervention to slow or prevent progression to cardio-metabolic disease.

Non-invasive sampling, such as exhaled breath, provides metabolic information that may be indicative of an individual’s health status. Exhaled breath metabolites have been reported in the assessment of infection^14,15^, cognition^16^, metabolic disease^17^, and lifestyle^18^. These studies provide evidence that metabolic markers, which strongly correlate with metabolic dysregulation, can be identified in breath samples. Previously, we reported an exhaled breath metabolomics study investigating a young cohort of non-human primates (NHP) who developed insulin insensitivity due to developmental programing^19^. Following the development of IR, a volatile organic compound (VOC) breath signature was identified that discriminated between NHPs with IR compared to controls. It is suggested the altered breath signature arose from an altered cardiometabolic state potentially due to increased ROS production, beta-oxidation, inflammation, and lipid peroxidation^19^. Based on these findings, we hypothesized that metabolic markers contained in exhaled breath could be a potential diagnostic resource for defining metabolic status in humans and furthermore could be used to diagnose IR in pre-diabetic adolescents. Our goals were to show feasibility of adolescent breath collections using the ReCIVA breath collection device (Owlstone medical, Cambridge, UK) and to identify a small set of breath biomarkers in a pre-diabetic population that are important for the detection of IR and the risks for cardio-metabolic disease development.

## 2. MATERIALS AND METHODS

### Ethical Approval of Study

All study procedures were approved by the Institutional Review Board for Baylor College of Medicine and Affiliated Hospitals (IRB-H-40940). Informed consent was obtained from parents or guardians and assent from participants. This work was performed in accordance with The Code of Ethics of the World Medical Association (Declaration of Helsinki) for experiments involving humans.

### Study design and participants

Participants were recruited from the Pediatric Health and Weight Management Clinic at the Children’s Hospital of San Antonio, TX. This cross-sectional study recruited Hispanic adolescents during their initial clinical visit. Criteria for inclusion were male and female ages ranging from 13-17 years with a BMI ≥ of the 95^th^ percentile. Exclusion criteria of the patients included a diagnosis of T2DM, asthma, or other chronic respiratory condition and any use of neurohormonal medications.

### Clinical Assessments

Upon arrival to the clinic, a trained medical technologist collected anthropometric measures. Height and weight were collected using a BSM170 digital stadiometer and Scale 570 (InBody, Cerritos, CA). BMI was calculated as Kg/m^2^ and z score calculated for all patients using their sex, precise age based on the date of birth and date of assessment, height, and weight. BMI percentile is expressed as the percent of the 95^th^ percentile. Participants and parents/guardians verified fasting status prior to blood samples being collected as a standard of clinical care blood work. Blood (3 mL) was collected in red-top vacutainer tubes (BD, Franklin Lakes, NJ). Clinical blood measures included: fasting insulin (mU/L) and fasting glucose (mg/dL), Hemoglobin A1c (HbA1c), alanine aminotransferase test (ALT), aspartate aminotransferase test (AST), fasting lipid profile, and high sensitivity CRP. The Homeostatic Model Assessment of Insulin Resistance, (HOMA-IR) of the participants was calculated based on the formula: fasting insulin (microU/L) × fasting glucose (nmol/L)/22.5^20^. The HOMA-IR categories are based on an adult Hispanic population based study^21^. **Table 1** reports the demographic and clinical measurement of all participants involved in this study.

**Table 1.**
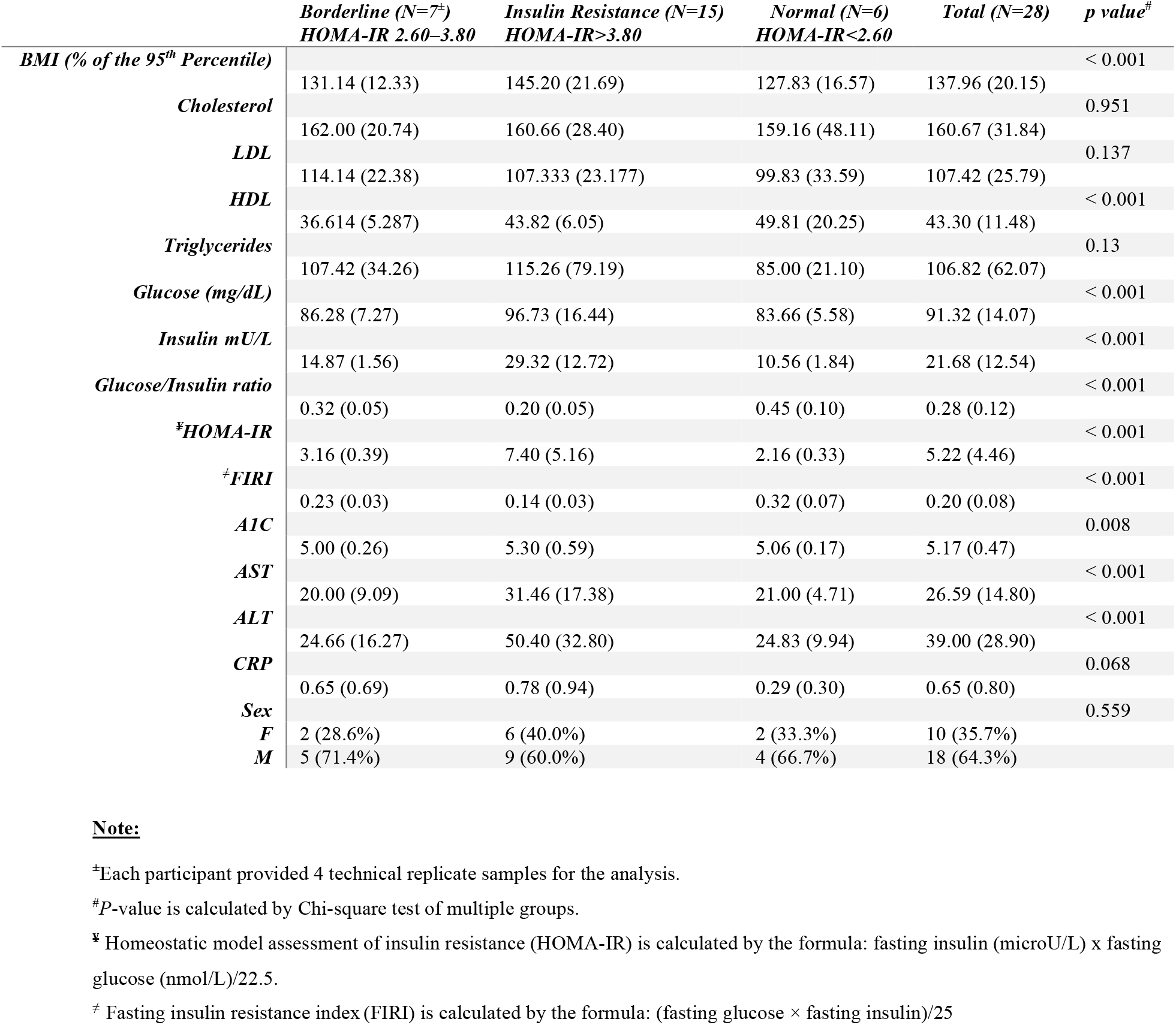
Demographic and clinical characteristics of the study cohort. The HOMA-IR cut-offs, are based on a study in a Hispanic population^21^. Data are presented as mean and standard deviation (SD).

### Exhaled Breath Sample Collection

On the same day of blood collection, participants provided exhaled breath samples through the breath collection protocol described below. The exhaled breath samples were collected using a commercially available breath sampler device, ReCIVA. This device safely monitors exhaled air pressure and CO_2_ production allowing for consistent exhaled breath collections. The ReCIVA device was supplied medical-grade air (Airgas, Radnor, PA) at a rate of 40 L/min per manufacture recommendations to avoid artifacts and contamination from room air. Exhaled breath metabolites were concentrated onto bio-monitoring thermal desorption tubes (TDT) (CAT# C2-CAXX-5149, Markes International, Sacramento, CA) designed to capture VOCs from breath. Exhaled breath was collected with the following protocol:

1. Device collection parameters were set prior to starting of the collection in ReCIVA Breath Sampling Controller Software (Owlstone medical, Cambridge, UK). Total collection volume was set to 500 mL per tube at a rate of 200 mL/ min for lower and upper airways.
2. Upon entering the participant room, a trained research assistant opened a new one-time use breath collection mask and assembled the four TDT into the ReCIVA device. The ReCIVA mask was placed over the participant’s mouth and nose and they were advised to breathe normally and familiarize themselves prior to the start of collection.
3. Once the participant acknowledged readiness, the breath collection was started by the research assistant.
4. Once the software acknowledged the collection was complete, the ReCIVA mask was removed from the participant. TDTs were capped, transported to the research laboratory at the Texas Biomedical Research Institute and stored at 4 °C until analysis.

### Thermal desorption and Analytical Instrumentation

The exhaled breath VOCs collected on the TDT were processed within 1 month of collection. TDT was desorbed at 260 ^◦^C for 10 minutes to a cryotrap (general-purpose CAT#U-T11GPC-2S) using a thermal desorption unit, TD100xr (Markes Int., Sacramento, CA). VOCs concentrated on the cryotrap were rapidly heated at 40 ^◦^C/min to 280 ^◦^C and desorbed to a comprehensive two-dimensional gas chromatography tandem Time-of-Flight mass spectrometer (GC×GC-TOFMS. LECO Corp, St. Joseph MI, USA). The column set was configured as follows: ^1^D Rxi-5Sil-MS (95% polydimethylsiloxane, 5% phenyl; 30 m × 0.25 mm dc, 0.25 μm d_f_) coupled with ^2^D Stabilwax column (Crossbond polyethylene glycol; 2.21 m × 0.18 mm dc, 0.18 μm d_f_)(Restek, Bellefonte PA, USA). The inlet and ^1^D column were connected by a PressFit column connector and the ^1^D and ^2^D were connected by an MXT-union connector kits (Restek, Bellefonte PA, USA). The ^2^D column end is feed into the transfer line to the MS. The helium carrier gas flow was set to 2 mL/min for the entire run. The GC oven temperature program was set to ramp from 40 °C to 100 °C by 5 °C /min and then ramp by 8 °C /min to 220 °C with a final 10 minute hold for a total run time of 38 minutes.

### Spectral data analysis

Raw data were processed using Chromatof software (version 4.72, LECO Corp, St. Joseph MI, USA). Spectra were collected over the range of m/z 40-400 at a rate of 100Hz. For peak findings, a signal-to-noise (S/N) cutoff was set to 100 and the NIST 2020 library was used for the identification of the analytes with a cut off of 700 match similarity. Analytes were aligned across all participants using Statistical-Compare tool contained within the Chromatof software. A chemical name was assigned if the analytes matched the following four criteria, 1) high mass spectral match, 2) group separation based on the structural formula 3) Retention Index match and 4) the extracted ion chromatogram (EIC) ionization patterns among all observed samples^22^. An analyte was not given a name if it did not match any of the four criteria. Possible contaminants were manually removed before further data analysis (**Supplementary Table S2**). Aligned data were exported for further data analysis.

### Data availability statement

Raw spectral data will be made available upon request.

### Data reduction and normalization

A brief summary of our data cleaning and feature reduction process is shown in **Supplementary Figure S1**. All statistical analyses were conducted in R 4.0.3 (R Core Team, Vienna, Austria)^23^. Data cleaning was performed as described^14,15^. In short, a frequency of observation (FOO) cut-offs of 80% was implemented to remove sparse features. The remaining features were normalized using Probabilistic Quotient Normalization (PQN)^24^, log_10_ transformed, and mean-centered. Missing values were imputed using a half-minimum approach^25^.

### Feature Selection, Machine learning and Statistical Analysis

A Random Forest (RF) regression model was implemented to select features clustering between individuals with different IR status. The important features were selected by considering all four samples collected from each participant. Combining male (n=18) and female (n=10) samples, a total of 112 (n=28×4) samples were analyzed. The samples were then divided randomly, 3:1, for a training and validation dataset.

The RF model was built using a 10-fold cross-validation (CV) scheme in the ‘caret’ package^26^. CV split the data into ten equal size pieces, building a model on nine of the ten pieces, and testing on the one remaining piece. The algorithm then leaves a different piece out and repeats this process for all pieces allowing for parameter tuning across the model, as well as estimating the model’s generalizability by examining accuracy statistics across the left-out pieces. Features were selected by an elbow cutoff of the RF variables important measure. The variables were given an importance measure by mean decrease accuracy method, which leaves one variable and builds a model with the rest of the variables.

Orthogonal partial least-squares discriminant analysis (OPLS-DA) was performed using the “ropls” package ^27^. Network analysis was performed in the MetScape 3^28^ for Cytoscape^29^ platform using the Kyoto Encyclopedia of Genes and Genomes (KEGG) ID and Human Metabolome Database (HMDB) ID. Additional statistical analysis was generated using Arsenal^33^ package of R^23^. For Pearson correlations, data were log_10_-transformed and correlations were conducted in R^23^ using ‘ggpubr’^35^. Boxplots were generated using the Wilcoxon non-parametric test, performed comparing each IR group type. Figure 3 created in R^23^ using ‘ggplot2’^37^ and ‘ggpubr’^35^. The ROC curve is generated in R^23^ using the ‘MLeval’^40^.

Biorender was used for generating Figure 6^30^.

## 3. RESULTS

### 3.1 Feasibility of breath sampling from prediabetic adolescents

Twenty-eight adolescents participated in this study with a mean age of 15.46 years old (range 13-17 years old). All participants were obese or severely obese with an average BMI percent of the 95^th^ percentile of 137.96 ± 20.15 (range 95-186), which expresses the extent of obesity (BMI ≥ the 95^th^ percentile are considered obese) among those adolescents^31^. The average exhaled breath sample collection time was 4.10 ± 0.05 minutes. At the time of collection all participants had not developed T2DM based on clinical blood measurements and clinician assessment.

### 3.2 Increased cardiovascular risk factors in Hispanic adolescents with obesity

As part of the routine clinical assessment, clinical blood tests were performed, and clinical characteristic are included in **Table 1**. Fasting glucose and insulin measures were used to calculate HOMA-IR, a surrogate measure of insulin sensitivity^32^. Based on HOMA-IR measures, three groups could be defined in our cohort, adolescents without IR (normal), adolescents with IR and adolescents with borderline IR with a statistically significant difference in IR value at < 0.001 in a Chi-square test of multiple groups. Adolescents with IR had a statistically significant higher BMI percentile (*p* < 0.001) with an average of 145.20 ± 21.69 compared with the normal adolescents 127.83 ± 16.57. Further emphasizing a prediabetic status of the cohort, measures of cardiovascular disease (CVD) risk factors, were normal for total cholesterol (average 160.67 ± 31.84 mg/dL) and LDL-cholesterol (average 107.42 ± 25.79 mg/dL) and low for HDL-cholesterol (43.30 ± 11.48 mg/dL). However, normal fasting glucose (91.32 ± 14.07 mg/d) and fasting insulin (21.68 ± 12.54 mU/L) levels were significantly different (p< 0.001) in adolescents with IR. Detailed clinical measures are presented in **Table 1**.

### 3.3 Machine learning approach identifies important breathprint for IR

The RF regression model identified the most important breath features for identifying individuals with IR based on their HOMA-IR value. Ten features were selected with the highest importance as shown on the RF importance plot (**Supplementary Figure S2)**. The elbow cut-off >18 were selected based on the large drop on the important measures on the mean decrease in Gini in the RF model^34^.

The ten important analytes include limonene, decane-2,4,6-trimethyl, undecane, undecane-2,7-dimethyl, pentylbenzene, octamethyloctane, eicosane and three unknown analytes (**Supplementary Table S1**). Identification of these analytes was confirmed by retention indices and mass spectral match score with NIST 2020 library’s analytical standard (level 2 and 3 identification)^22^. The unknowns reported are those whose retention indices were consistent, but the mass spectra matching was inconsistent or below 700 similarity score, hence a compound name was not assigned. Chromatographic and mass spectral information on the ten compounds is provided in the **Supplementary Table S1**. An example of a peak detection and identification used in this study are shown in **Supplementary Figure S3**.

### 3.4 The breath-IR model correlates with the standard blood based HOMA-IR

The ten breath VOCs identified in the training dataset, were evaluated against the remaining samples in the test dataset (**Figure 1a**). Test dataset comprised of a total of 37 samples that were not used for the training dataset. The test dataset also resulted in a positive correlation with blood based HOMA-IR values with a Pearson correlation, R=0.87, *p* <0.001. Combing the training and test datasets in a total dataset resulted in a higher correlation with the blood based HOMA-IR values with a Pearson correlation, R = 0.95, and *p* <0.001 (**Figure 1b**).

**Figure 1:**
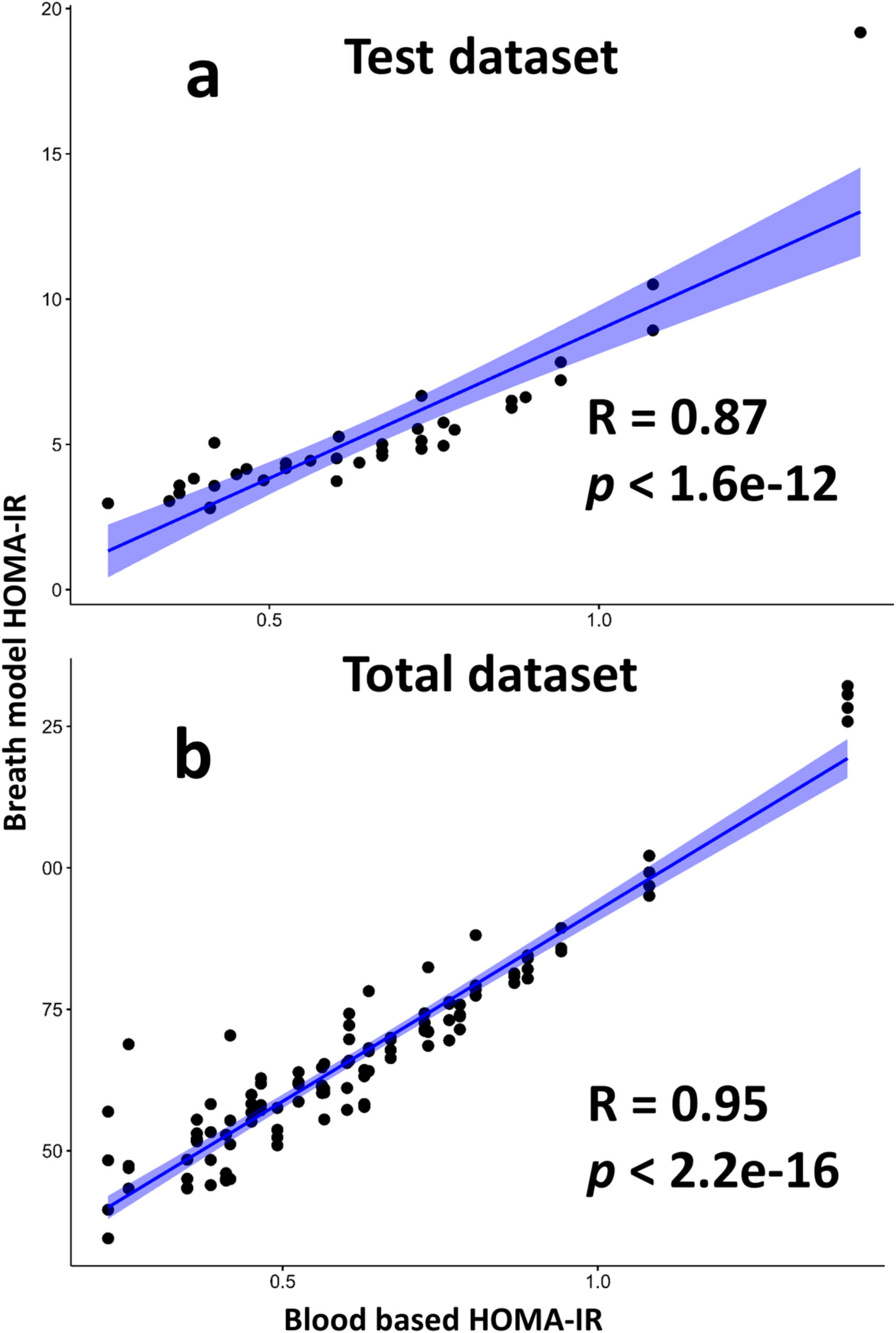
Pearson correlation of the breath-based IR with the blood based HOMA-IR measurement. (a) Correlation of a test dataset against the blood based HOMA-IR measures testing the Breath IR training set on independent samples. (b) Correlation of the total dataset combining training and test datasets for the breath-IR model against blood based HOMA-IR.

### 3.5 Breath-IR model correlation with the glycemic profile but not with the lipid profile

The breath IR model shows a strong correlation with measures of glucose metabolism including fasting glucose (mg/dL) and fasting insulin (mU/L) with R=0.91, *p* < 0.001 and R=0.80, *p* <0.001, respectively **(Figure 2a and b)**. A weak positive correlation was observed with triglycerides (mg/dL), R=0.35, *p* <0.001 and weak negative but non-significant correlation with the LDL levels at R=-0.028, *p* <0.77 (**Figure 2c and d)**. The breath IR model did not correlate with the total cholesterol (mg/dL) or HDL level mg/dL **Supplementary Figure S4**. The breath-IR model also correlated with additional indices of IR; fasting insulin resistance index (FIRI)^36^ and the glucose/insulin (G/I) ratio showed in the **Supplementary Figure S4**. The HbA1c test positively correlated with the breath IR model, R = 0.63 and *p* = <0.001 (**Supplementary Figure S4)**.

**Figure 2:**
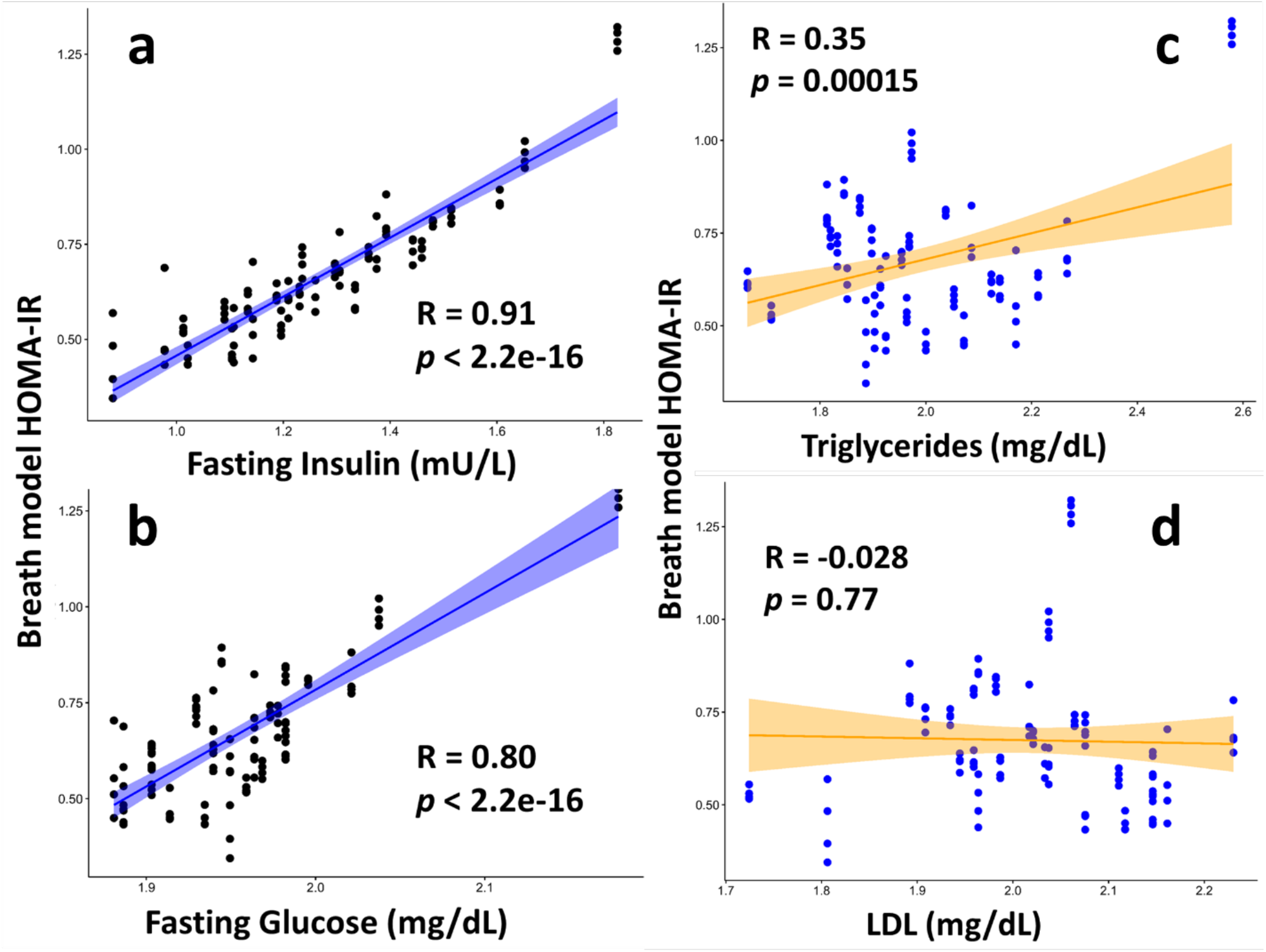
Pearson correlation of the breath based HOMA-IR with glycemic and lipid measures. Pearson correlation was performed to compare the breath based HOMA-IR model against (a) fasting insulin (mU/L), (b) fasting glucose (mg/dL), (c) triglycerides (mg/dL) and (d) LDL (mg/dL).

### 3.4 Breath-IR model accurately identifies the IR classification

To determine the breath-IR model accuracy for identifying individuals with IR, the ten features identified by RF were further investigated. Four of the ten VOCs, limonene, undecane, undecane, 2,7-dimethyl and unknown 1 were statistically significantly different for individuals with IR (*p* =0.003, *p* =0.002, *p*<0.001 and *p*=0.033, respectively) compared to those without IR (**Figure 3**). A trend was observed for the remaining analytes but was not statistically significant **Supplementary Figure S5**. A moderate difference was observed for limonene when comparing borderline IR individuals to normal (*p*=0.068), whereas comparison of borderline individuals to IR group was not significant (*p*=0.82).

**Figure 3:**
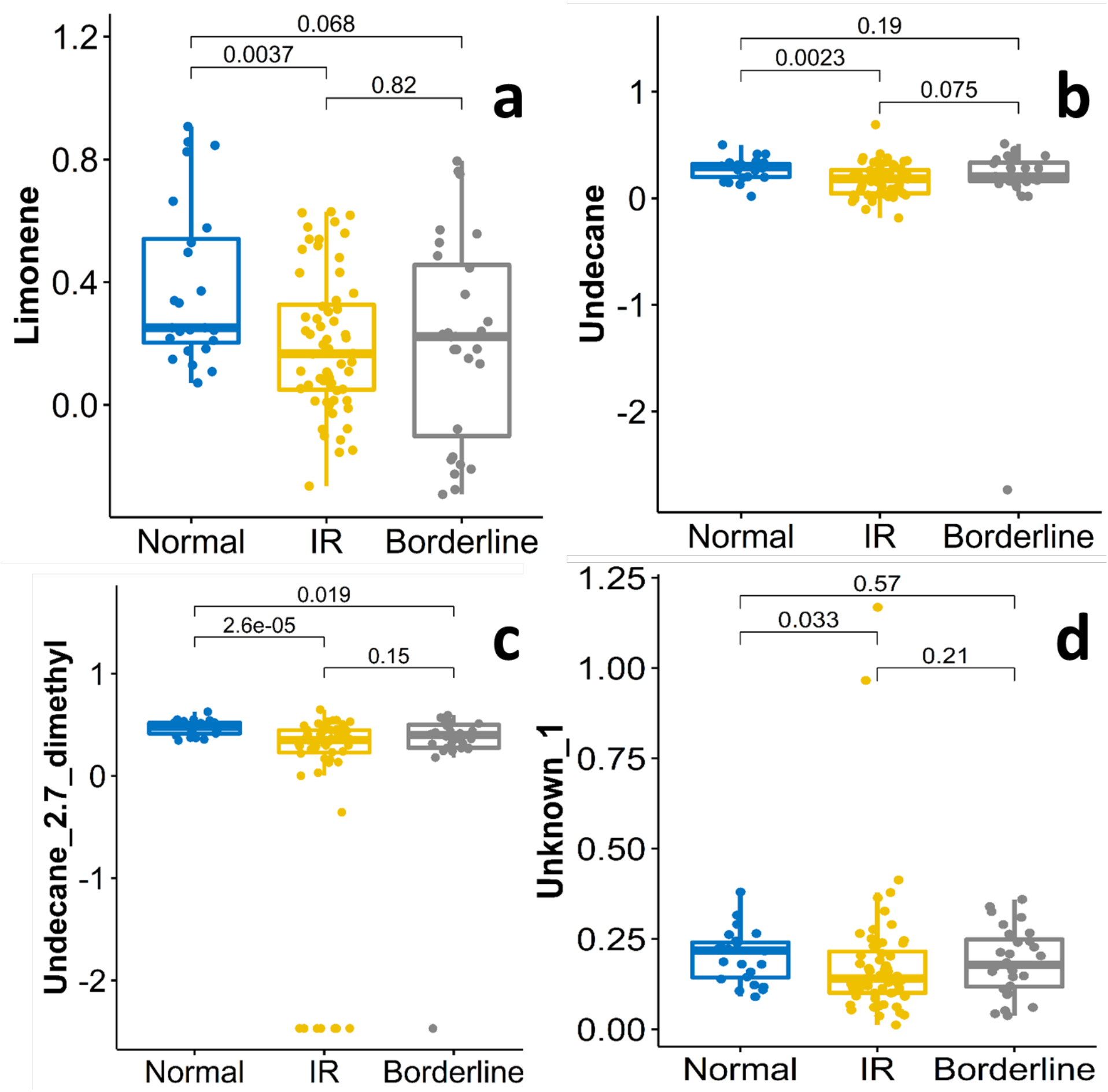
Boxplots of mean centered and normalized peak area for four compounds selected from the breath-IR model across normal, IR and borderline IR adolescents. Each compound’s median observed peak area between the groups were different indicating a univariate difference which may be contributing the discrimination of the IR group in the combined 10 compounds model. Boxplots represent the quartiles of the data (first line is the first quartile, midline is the median, third line is the third quartile) where whiskers represent 1.5 × IQR (inter-quartile range).

To evaluate the combined performance of the breath-IR model for clustering between the HOMA-IR categories, an Orthogonal Partial Least Squares Discriminatory Analysis (OPLS-DA) was performed^27,38,39^. The OPLS-DA showed a clear clustering by the HOMA-IR score, shown in **Figure 4**. Two extreme HOMA-IR values (24.92, 12.07) were removed from the dataset to show clearer clustering, but even with those extreme values, a similar pattern of clustering of the individuals based on the breath-IR model was observed (**Supplementary Figure S6**).

**Figure 4:**
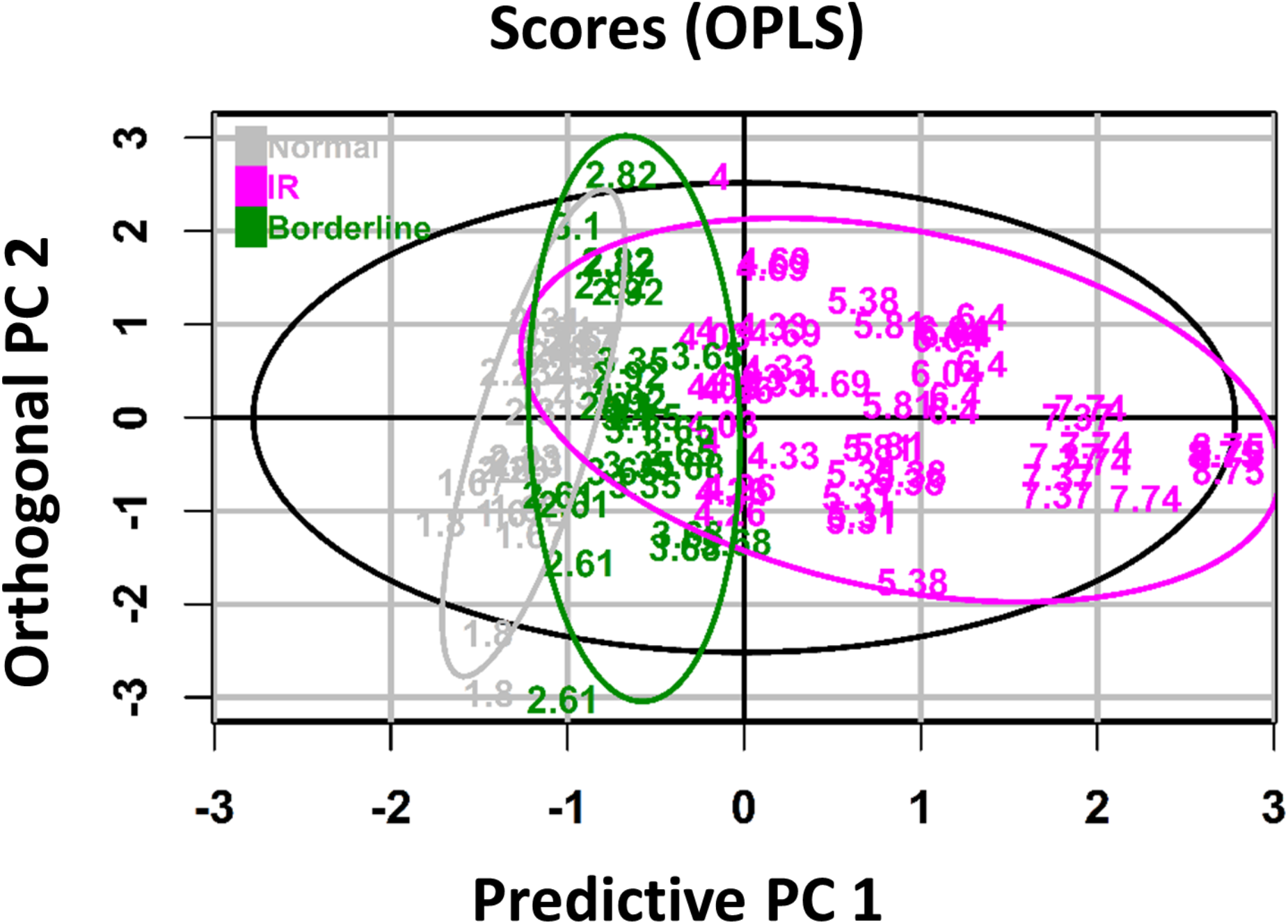
The supervised Orthogonal partial least-squares discriminant (OPLS) analysis of breath HOMA-IR model. The score scatter plot of OPLS model showing the clustering of samples in horizontally by predictive principal component (PC) 1 and vertically by orthogonal PC 2. The IR cut-off, is based on a study in Hispanic population^21^. The purple color indicates the IR group with HOMA-IR>3.80, the green color represents the borderline IR group with HOMA-IR 2.60–3.80 and HOMA-IR<2.60 indicate normal group which is presented as grey color. The 95% tolerance region corresponds to the ellipse that is defined by the Hotelling’s T2 parameter.

To evaluate the performance of the breath-IR model for classifying individuals with IR, the area under the receiver operating characteristic curve (AUC -ROC) was plotted resulting in a breath IR model value of 0.87 (**Figure 5**). This value is generated by an RF iteration based on the selected ten breath compounds. The process was repeated for only four VOCs that were significantly different for individuals with IR, limonene, undecane, undecane, 2,7-dimethyl and unknown 1 reducing to 0.83. The RF model’s importance was also assessed by randomly selecting features in the dataset resulting in a AUC of 0.57. The breath-IR model observed sensitivity and specificity as 73.1% (60.0 to 83.0% in cross-validation) and 81.7% (70.0 to 89.0% in cross-validation), respectively.

**Figure 5:**
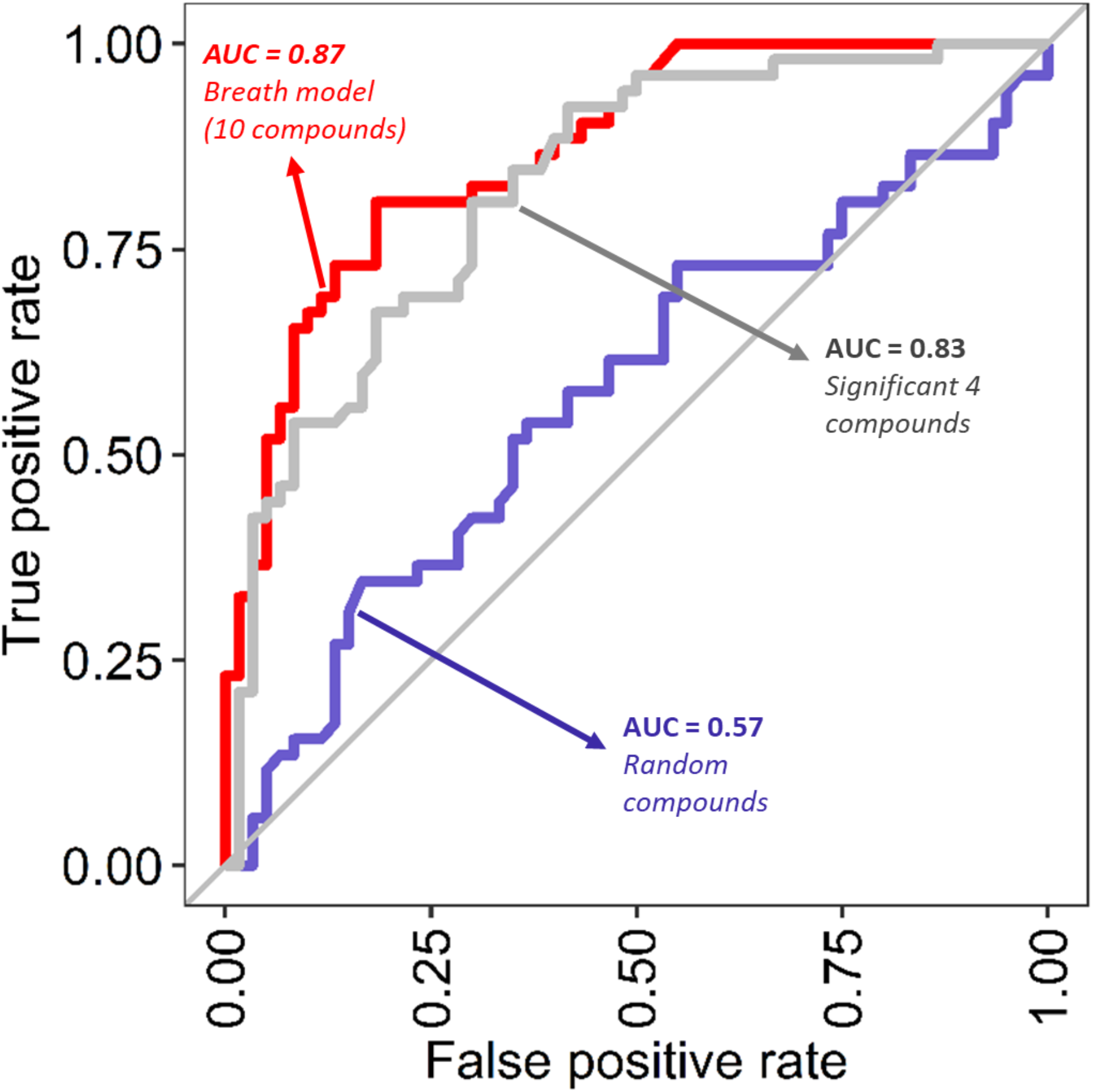
Diagnostics performance of the Breath-IR model. The receiver operating characters (ROC) curves using the random forest model between the IR (HOMA-IR>3.80) and normal (HOMA-IR ≤ 3.80) adolescents. The ‘red line’ indicates the model’s performance based on 10 compounds in breath IR model, the ‘grey line’ indicate performance by only four compounds with individual statistically significant difference, limonene, undecane, undecane, 2,7-dimethyl and unknown 1. The ‘blue line’ indicates randomly selected 4 breath metabolites from the original dataset. The AUC-ROC across folds after cross validation are 0.87 (0.80-0.94) for 10 features breath model, 0.83 (0.75-0.91) for significant 4 compounds, and 0.57 (0.46-0.68) for random features model.

**Figure 6:**
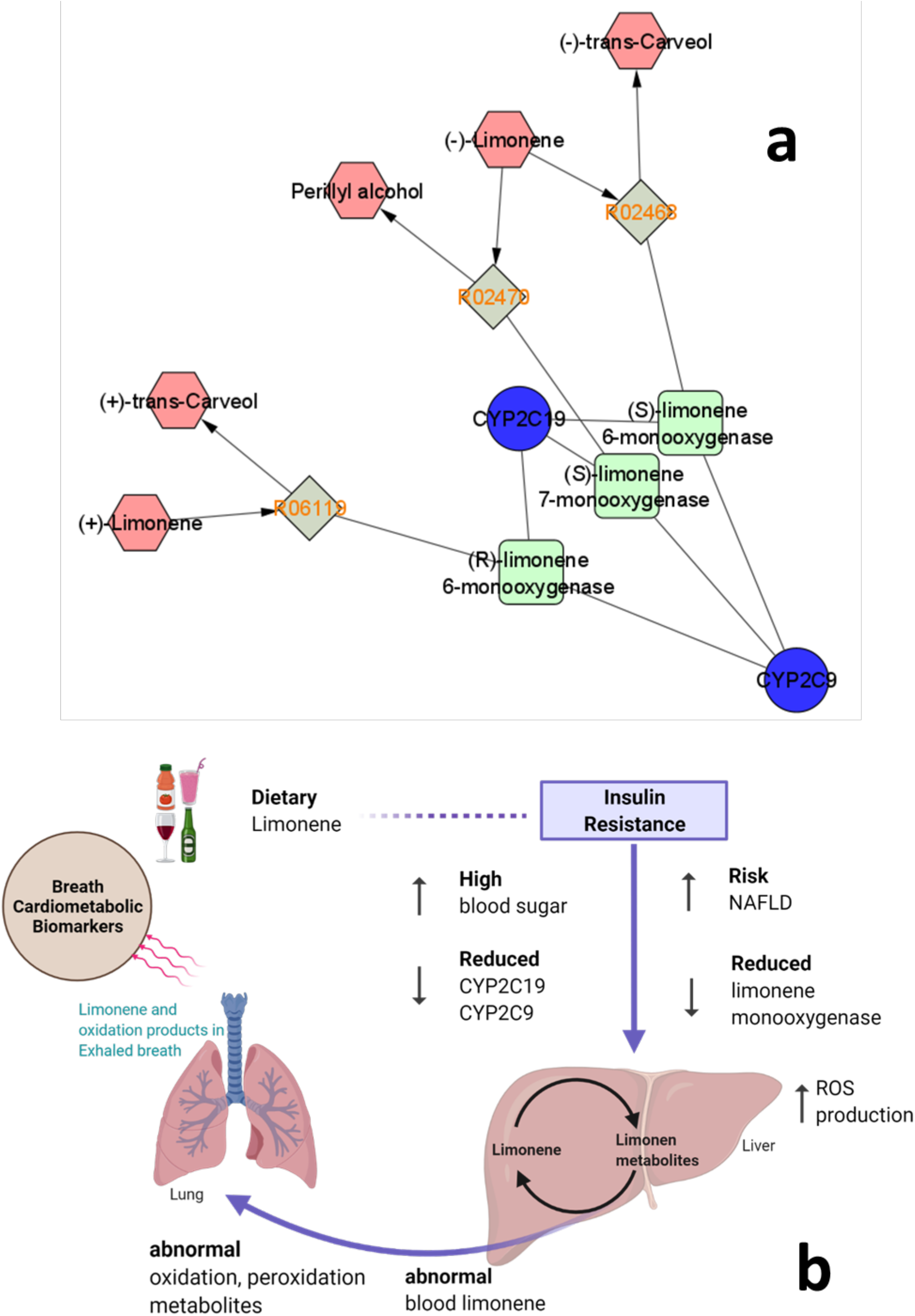
Molecular network analysis of limonene. The complex interaction of the gene, engymes and the metabolites are presented by Cytoscape^29^ platform using Metscape 2^28^ network anlaysis **(a)**. The reaction R02468, R02470 and R06119 connect the injested (+) limonene and (-) limonene to its oxidized metabolites, (+) trans-carveol, (-) trans-carveol and perillyl alcohol. The enzymes responsible for these reactions are P450 enzymes CYP2C9 and CYP2C19 and their products (S)-limonene 6-monooxygenase, (S)-limonene 7-monooxygenase and (R)-limonene 6-monooxygenase and **(b)** the proposed mechanism of the breath cardio-metabolic biomarkers. Increased ROS production, abnormal limonene and oxidation metabolites due to liver damage may lead to an abnormal level of breath metabolites. The figure is created using biorender^30^ platform.

## 4. DISCUSSION

### 4.1 Non-invasive exhaled breath as IR diagnosis

Our study shows the feasibility of collecting non-invasive exhaled breath metabolites from an adolescent population, which can then be used to assess an individual’s metabolic health status. Utilizing an untargeted metabolomics approach allowed for the identification of all breath metabolites in detectable range of our mass spectrometer. By implementing a machine learning model to assess the normalized data, a predictive model based on the selected features could be built to inform diagnostic potential of exhaled breath signatures. Our previous studies in nonhuman primates identified exhaled breath signatures in animals with IR^19^. The adoption of the HOMA-IR categories was based on a population-based study reported from the adult Mexican Americans population^21^, as no clear definition exist for the Hispanic adolescents with IR. Our breath-IR model strongly correlates with all glycemic clinical measurements, but the model did not strongly correlate other clinical measures (i.e., circulating lipids). Lipid levels were not in a range that would have clinically categorized the participants as having T2DM, therefore the breath IR model is specific to measures impacting IR. Given the breath-IR model resulted in a strong correlation with blood based HOMA-IR measurements, information contained in exhaled breath samples has utility for a non-invasive IR diagnostic.

This pilot study is exploratory in nature and our breath metabolites distribution is based on relative abundance of each VOC. Therefore, metabolite levels are based on the normalized peak area not the actual quantity of metabolites. Four of the ten important features identified in the breath-IR model limonene, undecane, undecane, 2,7-dimethyl and unknown 1 showed a statistically significate difference between the normal and IR group. These four metabolites are the major contributors to the accuracy of the model. The OPLS-DA model filtered out noise in the dataset that is not correlated with the outcome variables^41^. Given two of the adolescents had an exceptionally high HOMA-IR (24.92 and 12.07), these two values put a biased weight on the classification problem. Although those values are extreme within the current cohort, two different OPLS-DA models, one without those values and one with those values, as shown in **Figure 2** and **Supplementary Figure S6**, respectively. These plots show clear clustering of individuals by their IR levels based on the breath metabolites, as IR individuals with HOMA-IR (>3.80) were clustered on the right side of the plots whereas the normal HOMA-IR individual (<2.60) on the left. The border line IR group with HOMA-IR 3.8-2.6 were also clustered in-between of the IR and normal group although there is some overlap. Although the other six breath metabolites did not reach significance between the groups individually (**Supplementary Figure S5**), their contribution to the breath-IR model is evident when assessing the diagnostic capability of the model (**Figure 5**) to identify individuals with IR. A combined signature of breath metabolites is clearly more informative than using a single metabolite which may be indicative of some normal variability in metabolites within and between individuals^15,19,42^.

Our breath IR model shows good accuracy 77.8% with sensitivity of 73.1% (60%-83% within cross-validation) and specificity of 81% (70%-89% within cross-validation) (**Figure 5**). As a comparison, the American Diabetes Association (ADA) and International Expert Council of the ADA define the diabetes diagnostic criteria for hemoglobin A1c (HbA1c); sensitivity, 73.3%; specificity, 88.2%^43,44^. Although our study needs to be verified in a larger more diverse independent cohort, our results are promising for the breath-based IR diagnostic tool for monitoring IR and metabolic health, which could be developed further.

### 4.2 Biological insight of the breath biomarkers

Among the seven named breath biomarkers, limonene, undecane, pentylbenzene, and eicosane have been previously reported in the human metabolome according to the KEGG^45^ and HMDB^46^ databases. Undecane was previously detected as a biomarker of nonalcoholic fatty liver disease (NAFLD)^47^, asthma^48^, and gastrointestinal disease^49^. Pentylbenzen and eicosane are hydrocarbon molecules and subcellular component of membrane epithelium^50,51^. These two analytes were previously reported from human metabolomics studies but were never quantified^46^. Similar hydrocarbon metabolites were reported as oxidative stress markers of disease in exhaled breath related to infectious disease^15^, heart disease^52,53^, psychiatric disease^54^, and cancer^55^. This mechanism involves hydrocarbons being triggered or released in to the breath by ROS, a form of oxygen with a reactive electron from the by-product of oxidation metabolism in the mitochondria. These highly reactive molecules when released to the cytoplasm have the potential to damage DNA, protein, and other cellular metabolites, producing a range of small chain saturated or unsaturated hydrocarbon^56^. These hydrocarbons then have the potential to enter blood circulation and be detected in the breath as non-specific oxidative markers due to normal blood gas exchange in the lungs. IR has been established as a strong contributor to oxidative stress and is known to damage the vascular lining of cells, increasing the risk of atherosclerosis^57^. By identifying oxidative markers in exhaled breath from the obese adolescents that correlate with altered metabolic status within this cohort, suggests the underlying oxidative stress may be specific to the IR development.

Several studies have reported limonene from blood, feces, urine, saliva, and breath^58-62^. Limonene is a monocyclic monoterpenoid, with one isoprene chain. It is naturally abundant in nature and used in the flavor and fragrance of foods and drinks. In exhaled breath studies of liver cirrhosis patients elevated level of limonene were observed compare to the controls^63-65^. As an exogenous metabolite, limonene is ingested during dietary intake and then metabolized in the liver by P450 enzymes CYP2C9 and CYP2C19 and their enzyme products (S)-limonene 6-monooxygenase, (S)-limonene 7-monooxygenase and (R)-limonene 6-monooxygenase (**Figure 6a**). Limonene can then be metabolized to trans-carveol and perillyl alcohol^66^. It has been noted that in patients with liver disease, particularly NAFLD at any stage, have a reduced capacity to produce both CYP2C and CYP2C19 enzymes^67^. Reduced liver capacity to metabolize limonene results in abnormal levels in circulation and excretion leading to a difference on the limonene which can be identified in exhaled breath (**Figure 6b**).

ROS arises from vascular organs including heart cells^68^ or kidney cells^69^ leading to a higher systemic level of oxidation and peroxidation products. This may include aliphatic or aromatic hydrocarbon, straight-chain mono-or poly-unsaturated aldehyde, straight-chain -mono or -poly unsaturated carboxylic acids, ester, epoxides, MUFAs and PUFAs^70^. Higher levels in the blood may cross the blood–air barrier within the gas exchanging region of the lungs and exhaled through breath. Thus a set of metabolites produced from the multiple organs as a result of cardio-metabolic disease development could potentially lead to a set of breath based cardio-metabolic biomarkers as indicated in **Figure 6b**. Our detection of limonene as an important breath metabolite for determining IR status, may be due to early signs of NAFLD in obese adolescents’ breath used in this study.

Given that the abnormal levels of limonene are present in individuals with IR suggest potential NAFLD and liver damage, we investigated clinical measures of two liver specific enzymes aspartate aminotransferase (AST) and alanine aminotransferase (ALT) and systemic inflammation marker c-reactive protein (CRP). AST and ALT are both liver specific enzymes that become elevated in blood as a result of liver damage. Our breath-IR model was slightly negative but significantly correlated with the AST/ALT ratio (**Supplementary Figure S7**). The AST/ALT ratio less than <1 is suggestive of NAFLD/NASH whereas the score >2 is suggestive of alcoholic liver disease^71^. A negative correlation between breath IR and AST/ALT ratio thus indicates adolescents have early NAFLD/NASH. Evaluation of CRP levels were moderately positive and statistically significant correlated with our breath-IR model (**Supplementary Figure S7**). This confirms that the presence of systemic inflammation that could negatively impact multiple organ systems throughout the body including the liver of those adolescents.

## 4. CONCLUSION

This pilot study demonstrates the feasibility of studying breath in adolescents and the potential of non-invasive breath metabolites as an important tool to detect IR in a unique cohort of Hispanic pre-diabetic adolescents with obesity. The breath model developed by a machine learning-based approach showed a strong correlation with the surrogate blood test for IR that is currently used in the clinical environment. The breath IR model confidently clustered individuals with and without IR, showing a promising diagnostics performance of the breath metabolites as evident by the ROC-AUC = 0.87. The detection breath metabolites as important features for IR status may result from early liver damage or due to increased cellular damage from ROS production as a result of obesity and lifestyle. Future studies will focus on validating the usefulness of breath signatures for detection and monitoring of IR in a larger population of at-risk individuals.

## Data Availability

Raw spectral data will be made available upon request.

## Acknowledgements

We would like to acknowledge the Julee Carlton and Veronica Duran from the Health and Weight Management Clinic, Children’s Hospital of San Antonio, San Antonio, TX who help collect the clinical measurement and facilitate sample collection. We also acknowledge the Texas Biomedical Research Institute for providing instrumental support of this project.

## Author Contribution

Conceptualization: ACB, LAC, GMK SC

Data curation: ACB, SC

Formal Analysis: MSK

Funding acquisition: LAC

Methodology: MSK, ACB, SC, LAC

Project administration: ACB

Resources: LAC, ACB

Visualization: MSK

Writing – original draft: MSK

Writing – review & editing: ACB, LAC, GMK, SC

