## Supplementary figures and tables for "Breath biomarkers of insulin resistance in pre-diabetic Hispanic adolescents with obesity"

Andrew C. Bishop, Ph.D.

Department of Internal Medicine, Section on Molecular Medicine

Wake Forest School of Medicine, Winston-Salem, NC

E:

O: 336-713-7148

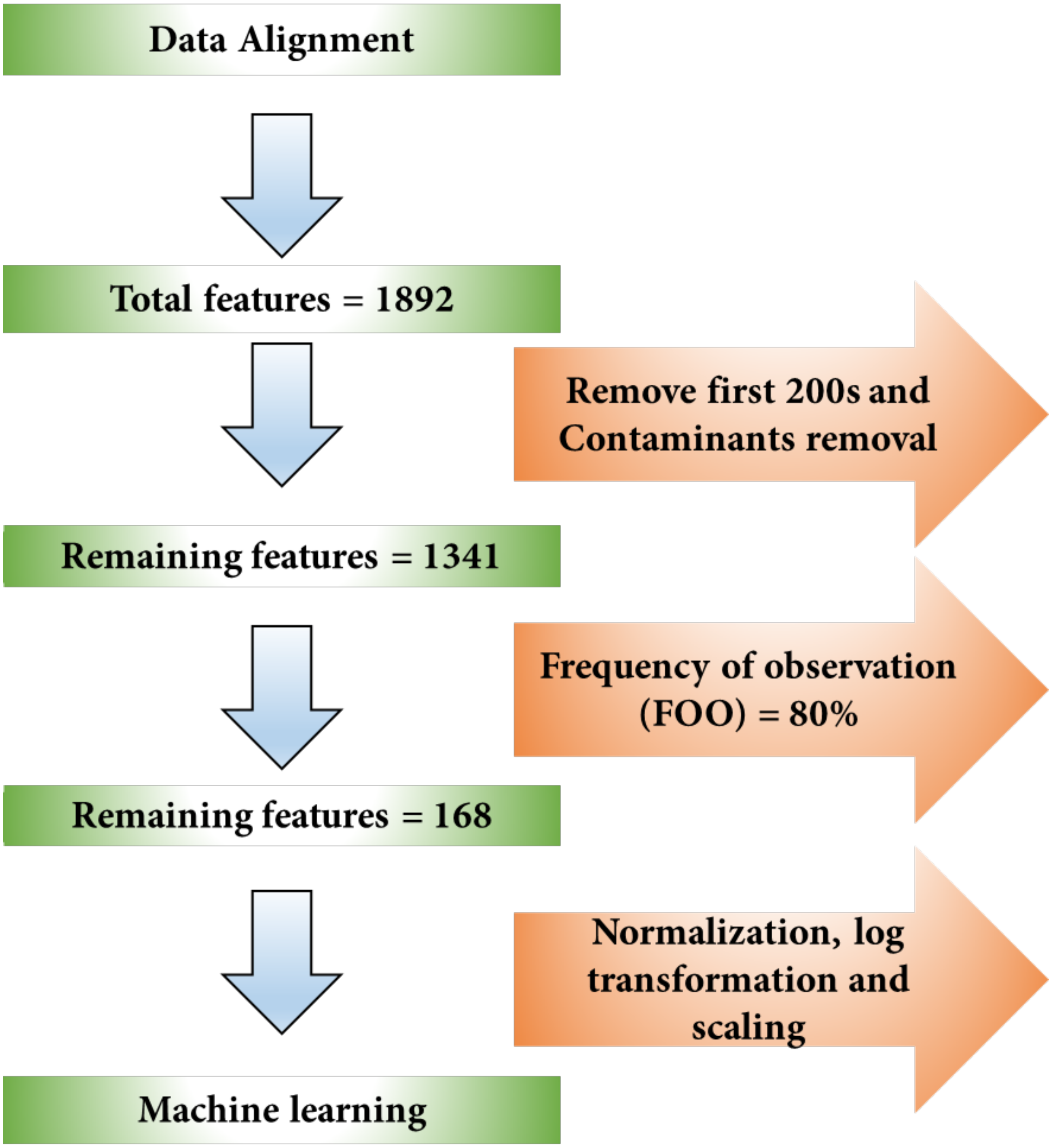


**Figure S1:** A flowchart of data analysis used in this study.

**Table S1:** Chromatographic and mass spectral identification of the ten compounds.

| Features | FINAL ID | Formula | RI[a] | Chemical Class | $^{1}t_{R}$(s) | $^{1}t_{R}$(s) (RSD %) | $^{2}t_{R}$(s) | $^{2}t_{R}$(s) (RSD %) | ID Matching[b] |
|---|---|---|---|---|---|---|---|---|---|
| 70 | Unknown 1 | | | | 564.3 | 0.3 | 0.2 | 30.0 | |
| 76 | Unknown 2 | | | | 594.3 | 0.3 | 1.3 | 1.9 | |
| 95 | Limonene | $C_{10}H_{16}$ | 1063 | cyclic monoterpene | 693.3 | 0.3 | 1.8 | 1.4 | RI + MS |
| 101 | Decane, 2,4,6-trimethyl- | $C_{13}H_{28}$ | 1095 | branched hydrocarbon | 745.0 | 0.0 | 1.4 | 1.6 | RI + MS |
| 105 | Undecane | $C_{11}H_{24}$ | 1141 | hydrocarbon | 824.4 | 0.2 | 1.4 | 1.6 | RI + MS |
| 111 | Undecane 2,7 dimethyl | $C_{13}H_{28}$ | 1201 | hydrocarbon | 915.2 | 0.1 | 1.4 | 1.7 | RI + MS |
| 112 | pentylbenzene | $C_{11}H_{16}$ | 1205 | aromatic hydrocarbon | 920.0 | 0.0 | 2.0 | 1.7 | RI + MS |
| 128 | Octamethyloctane | $C_{16}H_{34}$ | 1396 | branched hydrocarbon | 1153.2 | 0.6 | 1.4 | 1.9 | RI + MS |
| 155 | Unknown 3 | | | | 1563.8 | 0.1 | 1.5 | 3.3 | |
| 158 | Eicosane | $C_{20}H_{42}$ | 1938 | hydrocarbon | 1618.8 | 0.1 | 1.6 | 2.3 | RI + MS |

[a] Retention index was determined using $C_8$ ~ $C_{22}$ n-alkane standard solution.
[b] ID was matched by reported retention indices and mass-spectral matchings.

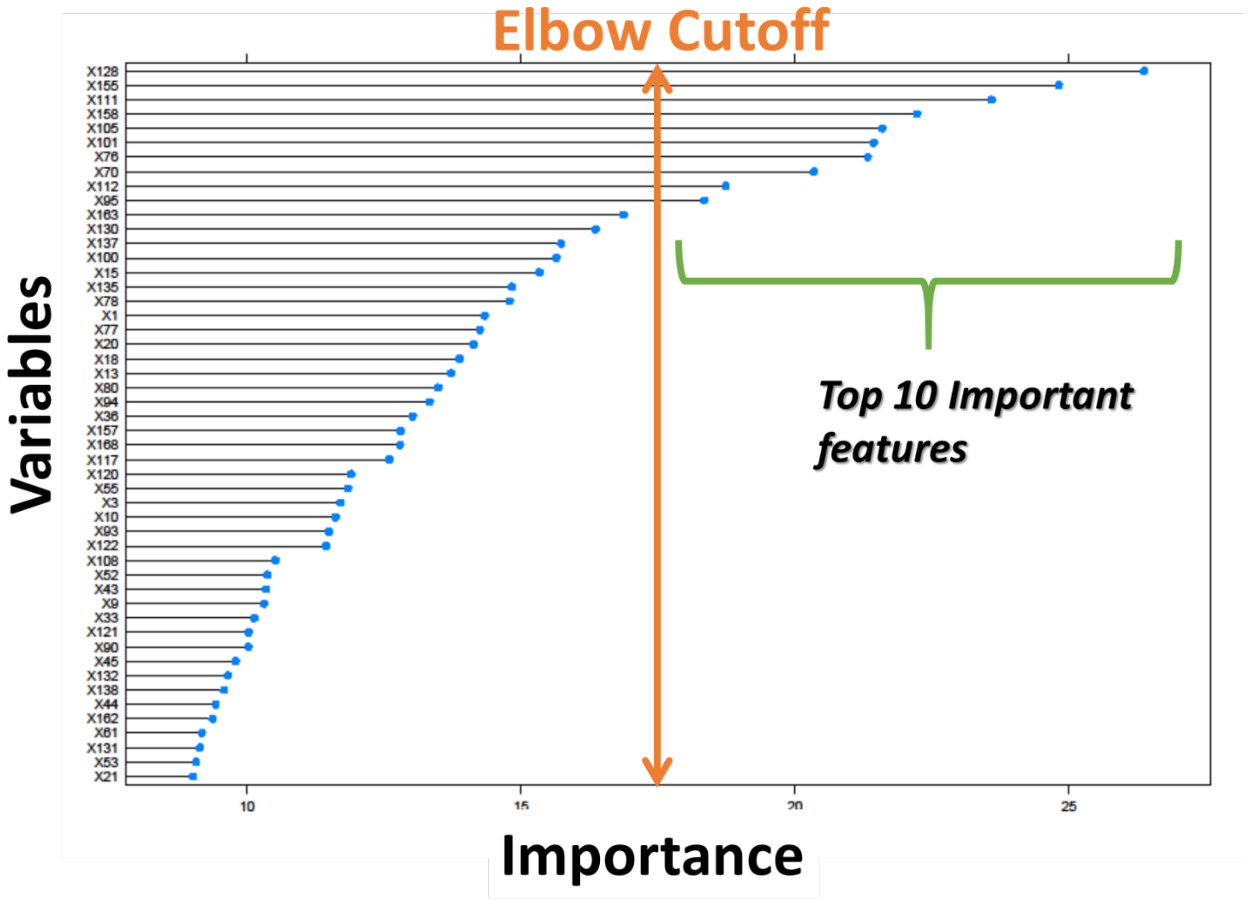


**Figure S2:** The importance plot of the Random Forest based feature selection. All variables are ranked based on the mean decease accuracy measure of Random forest. The elbow cutoff is based on the drop of importance which selected the top 10 important features for further analysis. The plot is generated in R[1] using the 'caret'[2] package.

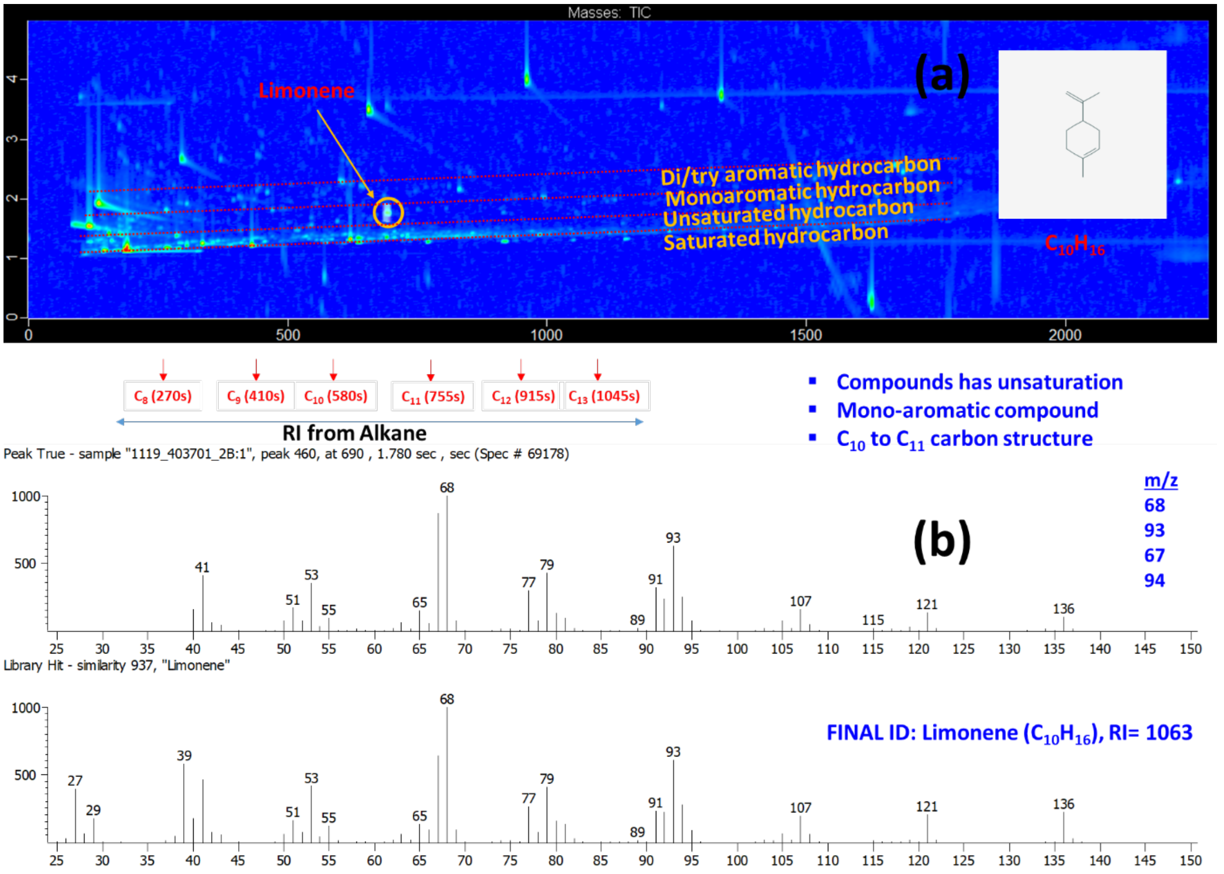


**Figure S3:** The example of a peak detection and identification used in this study. (a) The GC×GC contour plot of a breath sample. The group type separation indicate the peak is an unsaturated, mono terpene compounds. The RI of the peak indicate the position of the peak between $C_{10}$ to $C_{11}$. (b) The peak is then compared with the NIST library by m/z 68, 93, 67 and 94. Finally, combining all these info, a final ID was given as “limonene”. The GC×GC plot is generated in Chromatof software.

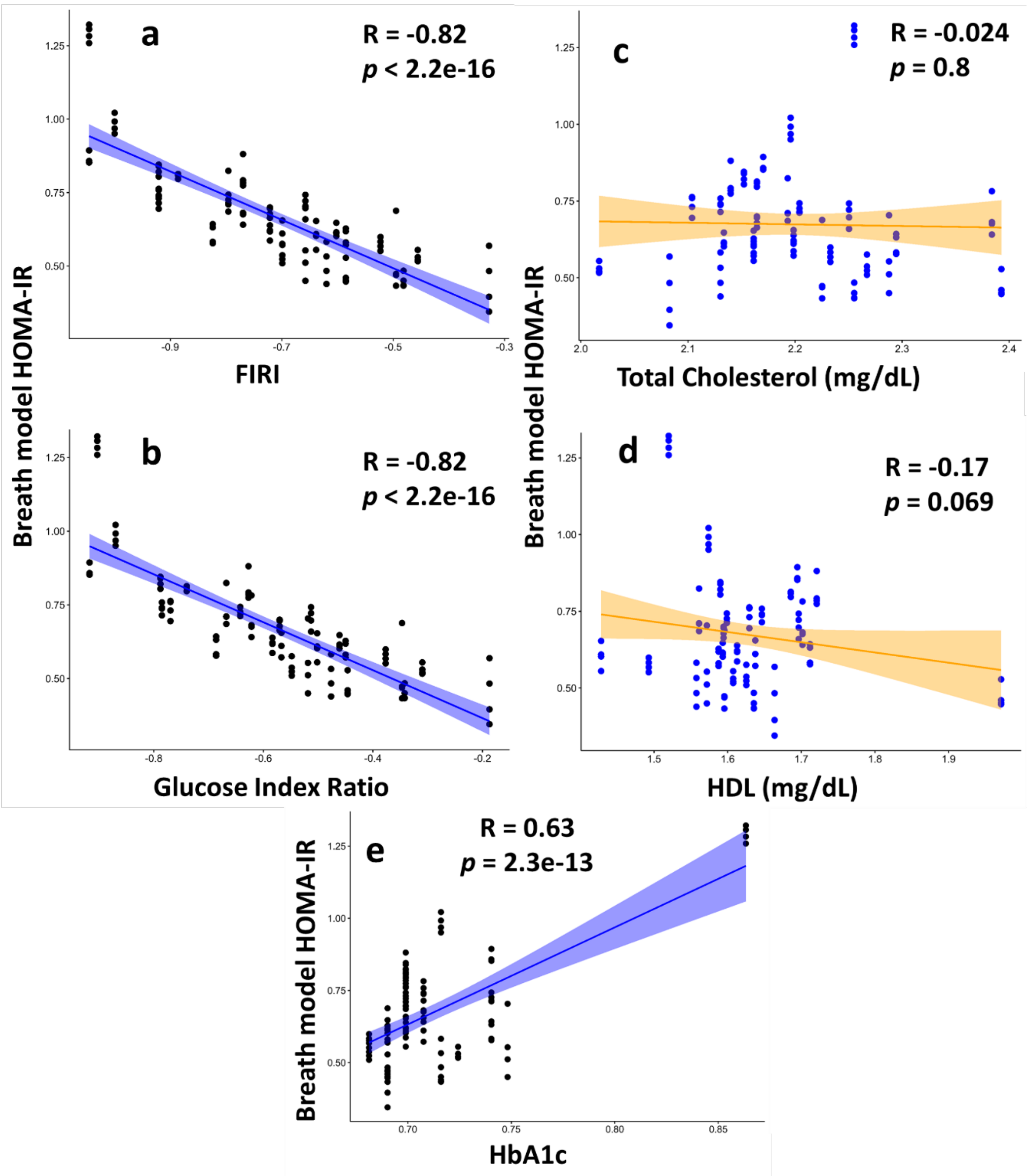


**Figure S4:** Correlation of breath based HOMA-IR model with Fasting insulin resistance index (a) (FIRI), (b) Glucose Insulin Ratio, (c) Total cholesterol (mg/dL), (d) HDL (mg/dL) and (e) HbA1c. Data were $\log_{10}$-transformed. Pearson correlation was conducted in R[1] using 'ggpubr'[3].

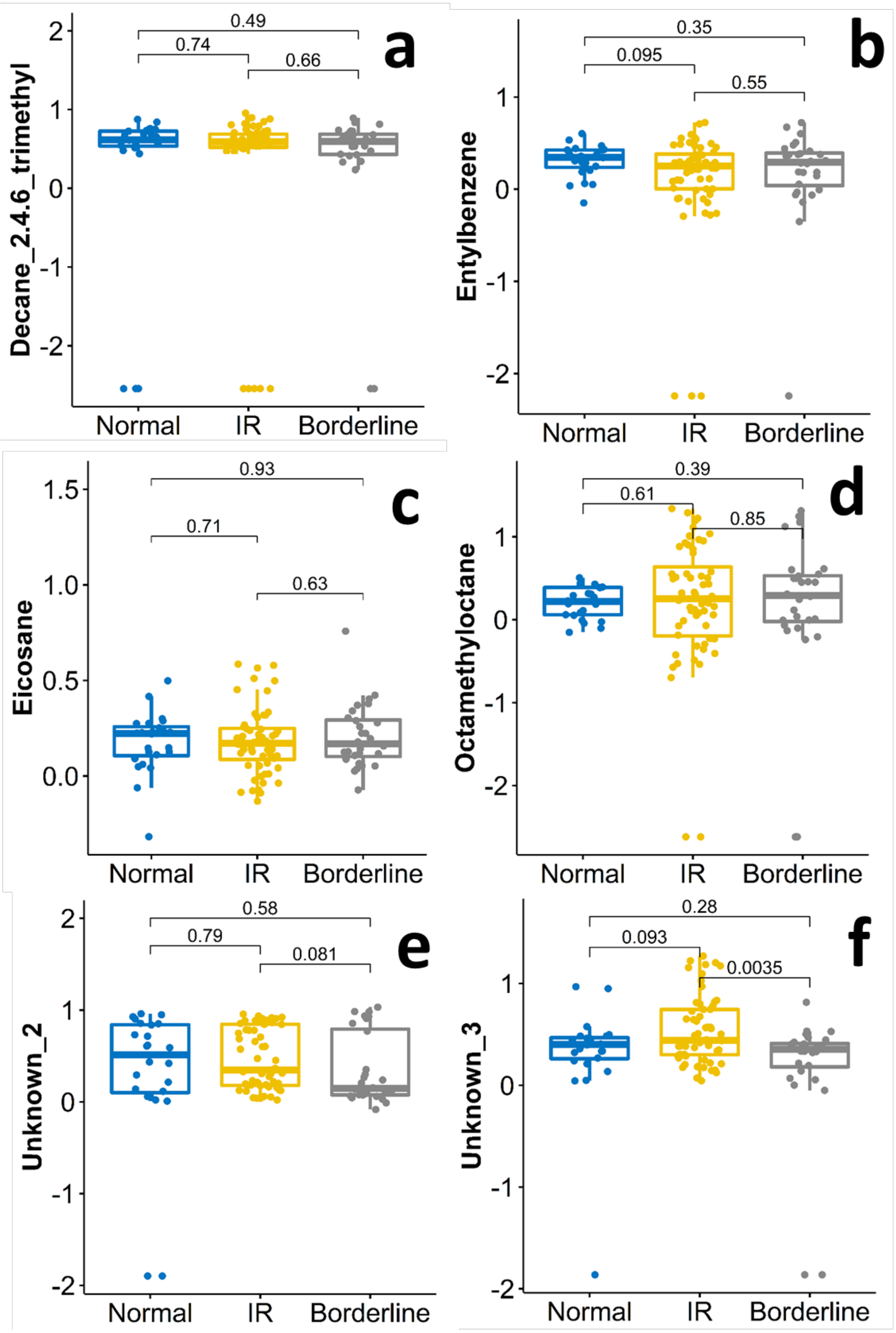
a
Decane_2.4.6_trimethyl
0.49
0.74
0.66
Normal
IR
Borderline
b
Entylbenzene
0.35
0.095
0.55
Normal
IR
Borderline
c
Eicosane
0.93
0.71
0.63
Normal
IR
Borderline
d
Octamethyloctane
0.39
0.61
0.85
Normal
IR
Borderline
e
Unknown_2
0.58
0.79
0.081
Normal
IR
Borderline
f
Unknown_3
0.28
0.093
0.0035
Normal
IR
Borderline

**Figure S5:** The boxplot of the mean centered and normalized peak area of (a) Decane_2.4.6_trimethyl, (b) Entylbenzene, (c) Eicosane, (d) Octamethyloctane (e) Unknown_2 and (f) Unknown_3. Boxplots show the quartiles of the data (first line is the first quartile, midline is the median, third line is the third quartile) where whiskers represent 1.5 × IQR (inter-quartile range). The Wilcoxon non-parametric test was performed comparing each IR group type. Figure created in R[1] using 'ggplot2'[4] and 'ggpubr'[3].

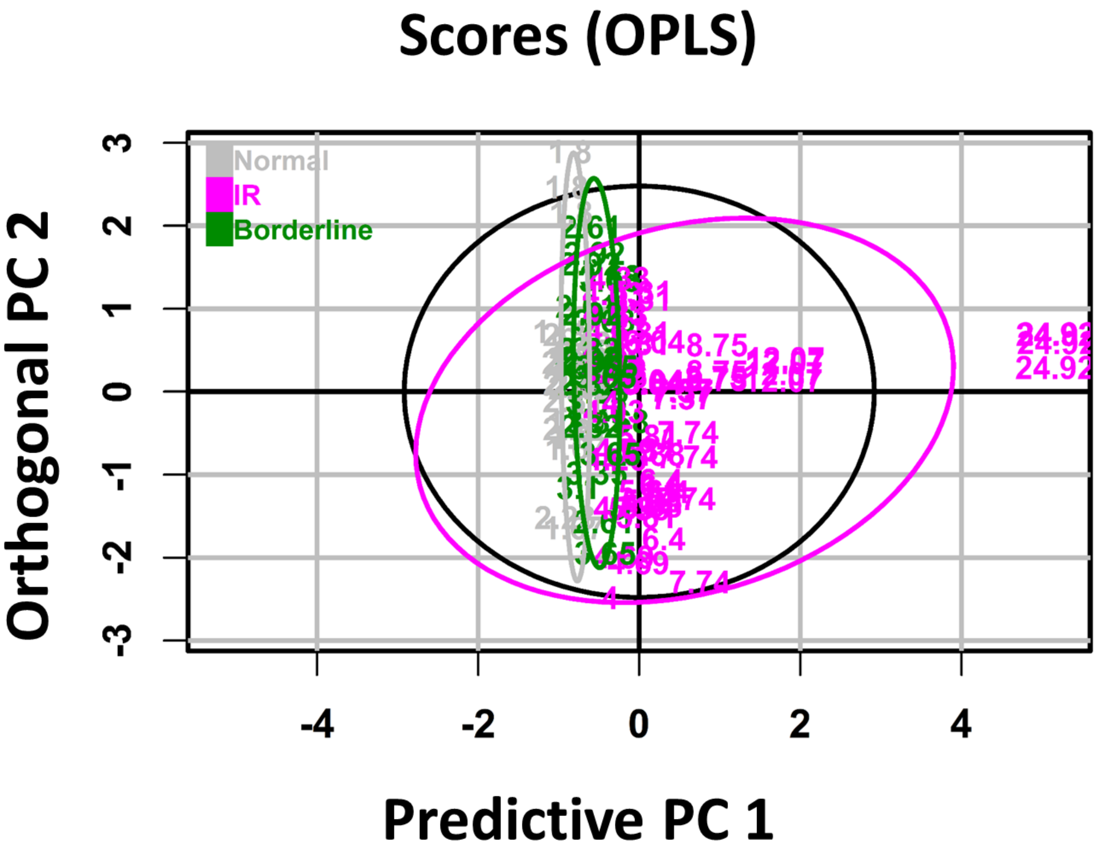


**Figure S6**: The orthogonal partial least-squares discriminant analysis (OPLS) plot of the IR samples with two extreme values. The purple color indicates the IR group with HOMA-IR>3.80, the green color represents the borderline group with HOMA-IR 2.60–3.80 and grey color indicates HOMA-IR<2.60 normal group. The 95% tolerance region corresponds to the ellipse that is defined by the Hotelling's T2 parameter. The OPLS analysis and figure created in R[1] using 'ropls'[5].

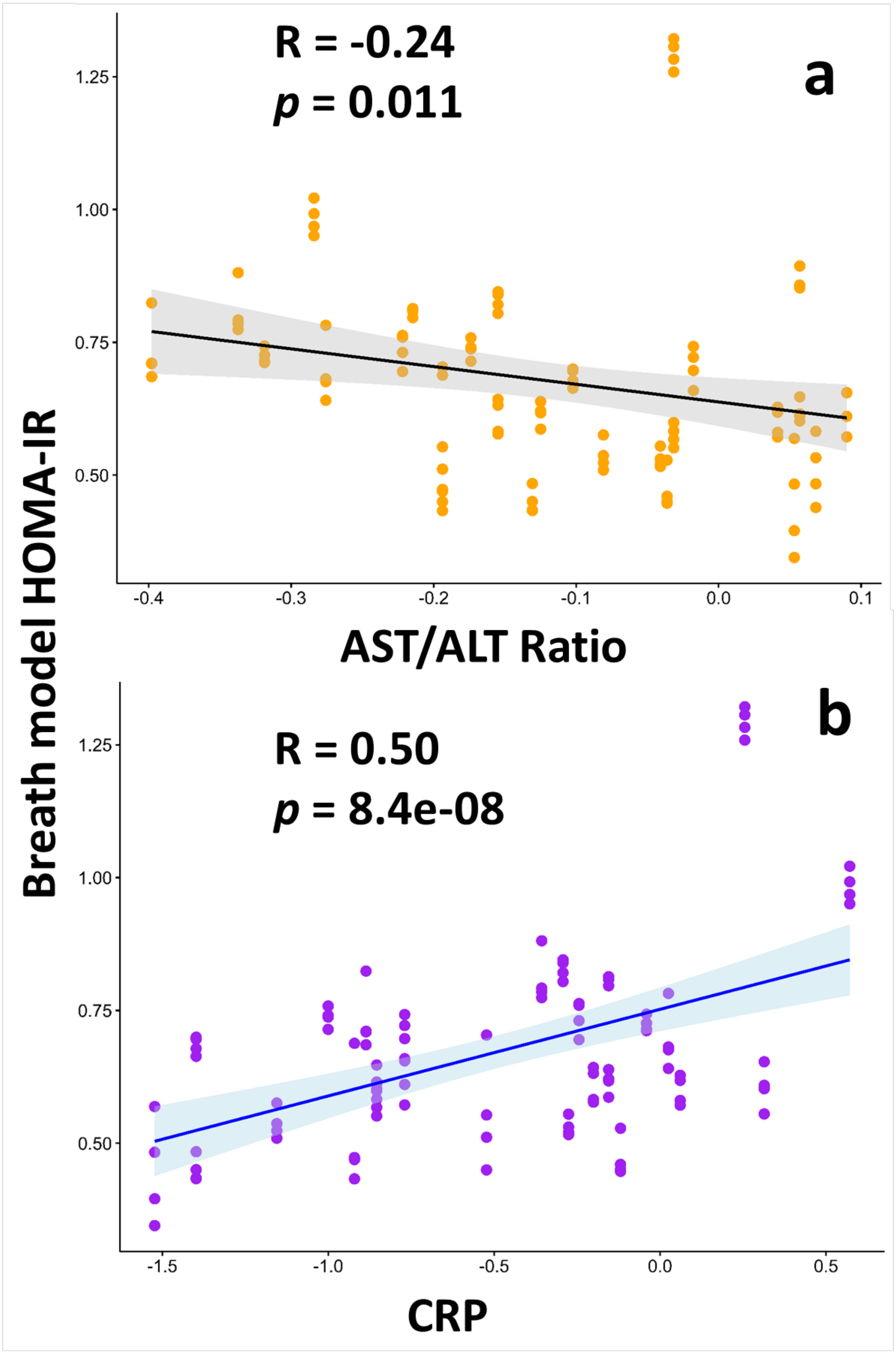
R = -0.24
p = 0.011
a
AST/ALT Ratio
R = 0.50
p = 8.4e-08
b
CRP
Breath model HOMA-IR

**Figure S7**: The correlation of the HOMA-IR of the adolescent, (a) the AST/ALT ratio and (b) c-reactive protein (CRP). Data were $log_{10}$ -transformed. Pearson correlation was conducted in R[1] using 'ggpubr'[3].

**Table S2:** List of known artifacts from the instrument and laboratory supplies used for the breath analysis are removed from the data analysis.

| **Contaminants Name** | **Frequency** |
|---|---|
| (2-(4-Isopropylphenyl)propan-2-ylperoxy)trimethylsilane | 2 |
| 1,1,1,3,5,5,5-Heptamethyltrisiloxane | 10 |
| 1,3,6-Trioxa-2-silacyclooctane, 2,2,-dimethylsilyl- | 2 |
| 1,3-Bis[methyl(trimethylene)silyloxy]propane | 1 |
| 1,3-Diallyl(tetramethyl)disiloxane | 1 |
| 1,3-Dioxa-2,4,6-trisilacyclohexane, 2,2,4,4,6,6-hexamethyl- | 1 |
| 1,4-Cyclohexadiene, 1,3,6-tris(trimethylsilyl)- | 1 |
| 2-Dimethyl(ethenyl)silyloxytetradecane | 1 |
| 2-Oxa-1,3-disilacyclohexane, 1,1,3,3-tetramethyl- | 1 |
| 4-Methyl-1-di(tert-butyl)silyloxypentane | 1 |
| Arsenous acid, tris(trimethylsilyl) ester | 1 |
| Carbamic acid, monoammonium salt | 17 |
| Carbon dioxide | 4 |
| Cyclotrisiloxane, hexamethyl- | 13 |
| Disiloxane, 1,3-diethyl-1,1,3,3,-tetramethyl- | 1 |
| Disiloxane, 1-ethenyl-1,1,3,3-tetramethyl-3-(2-propenyl)- | 4 |
| Disiloxane, ethylpentamethyl- | 4 |
| Disiloxane, hexamethyl- | 5 |
| Disiloxane, pentamethyl-2-propenyl- | 1 |
| Ethyl(dimethyl)isopropoxysilane | 1 |
| Methylene chloride | 2 |
| Nitrous oxide | 60 |
| Oxalic acid, 2TMS derivative | 4 |
| Oxalic acid, 6-ethyloct-3-yl isohexyl ester | 1 |
| Oxalic acid, butyl isobutyl ester | 1 |
| Oxalic acid, isobutyl nonyl ester | 1 |
| Oxalic acid, isobutyl pentyl ester | 1 |
| Phthalic acid, hex-3-yl isobutyl ester | 1 |
| Phthalic anhydride | 1 |
| Silane, diethoxydimethyl- | 1 |
| Silane, ethoxytriethyl- | 1 |
| Silane, methyldiisopropoxymethoxy- | 1 |
| Silane, methyltriisopropoxy- | 1 |
| Silane, tetramethyl- | 21 |
| Silane, trichlorodocosyl- | 1 |
| Silane, trimethyl-2-propenyl- | 1 |
| Silanediol, dimethyl-, diacetate | 1 |

| | |
|---|---|
| Silanol, trimethyl- | 4 |
| tert-Butyldimethylsilanol | 1 |
| tert-Butylpentamethyldisiloxane | 1 |
| Trimethyl(3,3-difluoro-2-propenyl)silane | 1 |
| Trimethylsilyl ethaneperoxoate | 1 |
| Trimethylsilyl-di(timethylsiloxy)-silane | 1 |
| Trisiloxane, octamethyl- | 12 |
